# Vaccination Behavior among Children with Developmental Delay and their siblings: A cross sectional study from Kerala

**DOI:** 10.1101/2020.05.11.20095026

**Authors:** Salah Basheer, NA Uvais

## Abstract

**Background and Aim:** Despite multiple scientific evidence to the contrary, parental concerns with respect to association of vaccination and development of Autism Spectrum Disorder (ASD) persist. Mental health professionals work closely with families with developmental disabilities. Greater understanding regarding the vaccination status in siblings of children with developmental disabilities and its associated factors will help them to contribute to public health strategies in combating infectious diseases. The aim of this study was to study the vaccination uptake and its associated factors in sibling of children with developmental disabilities.

**Materials and Methods:** The study design was cross sectional in nature. The families of children with developmental disabilities were recruited into the study from three child developmental centres. The data was collected using a semi-structured questionnaire. The recruitment of participants into the study was done from December 2017 to February 2018.

**Results:** 189 families with children with developmental disabilities were recruited into the study. In total, these children had 114 typically developing elder siblings and 50 typically developing younger sibling. The proportion of overall complete vaccination among children with developmental disabilities group and the younger sibling group were significantly lower than their older sibling group (P<0.01). The proportion of MMR vaccination among children with developmental disabilities group and the younger sibling group were significantly lower than their older sibling group (P<0.001).

**Conclusions:** Findings from this study suggest that the reduced vaccination uptake is a general trend in families of children with developmental delay. Such a significant fall in the vaccination rate in this group of children will make them vulnerable in case of outbreaks. This emphasise the need to have public health strategies targeted to improve the vaccination rate in families of children with developmental disabilities.

## Introduction

Vaccination hesitancy and vaccination refusal have been an issue since the practice of vaccination began. Recent outbreaks of various vaccination preventable diseases have emphasised the public health importance of this issue. In 2015 alone, more than three million children died globally, before the age of five, from preventable infectious disease.^[1]^ Identifying the factors affecting the vaccination behaviour is essential in order to effectively tackle this problem. In developing countries like India, lack of access to vaccination and family characteristics such as low education, literacy and socio-economic status make up the majority of reasons for poor vaccination among children.^[2]^

In addition to the attitudinal factors and religious beliefs, perceiving vaccination to cause adverse effects was consistently associated with vaccine refusal.^[3]^ Though, multiple studies have shown that there is no link between MMR (mumps, measles, and rubella) vaccine and development of Autism Spectrum Disorder (ASD), this perceived link has influenced public vaccination behaviour. This can be seen in previous studies that have shown that there is decreased vaccination uptake in younger siblings of children with ASD.^[4,5]^

However, there is a lack of studies that have explored whether this trend of decreased vaccination uptake is a general phenomenon in families of subjects with developmental disabilities. Thus, this study was aimed to compare the vaccination rate among children with developmental delay and their siblings.

## Methodology

The study design was cross sectional in nature and the protocol of the study was approved by the Institute Ethics Committee, Iqraa International Hospital and Research Centre, Kozhikode, Kerala. The families of children with developmental disabilities were recruited into the study from three child developmental centres in districts of Kozhikode and Kannur in Kerala, India. The data was collected using a semi-structured questionnaire by a trained therapist at each centre. The trained therapists who took part in the study included qualified speech therapist, physiotherapist and special educators. They were given information regarding the study and how to collect the data. These therapist were staffs of the centres were the children whose data was collected for this study were being given various therapies. The children who were participants of the study had been evaluated by qualified psychiatrists under whom the child is receiving treatment and the diagnosis was done as a part of routine clinical evaluation according to ICD 10 criteria. The diagnosis was recorded in the clinical files. The data regarding diagnosis was collected from these clinical files. No additional tools were used for diagnostic assessment. Though the current study involved a qualified psychiatrist, he was not directly involved in clinical diagnosis as a part of this study. Sample size was decided based previous similar studies.

Immunisation status was determined based on National Immunisation Schedule. MMR vaccination status at 18 months of age was also recorded. This parents who participated in the study were asked of they had given vaccination according to the national immunisation schedule. In addition, they were also asked whether they had given MMR vaccination. Informed consent was taken from the primary care giver of the child prior to enrolment into the study. The recruitment of participants into the study was done from December 2017 to February 2018.

This study was aimed to study whether there were changes in vaccination behaviour between siblings. The vaccination behaviour in normal elder and younger siblings was studied as a part of the study and normal elder siblings were conceptualised as a control group against which patients and their younger siblings were compared.

The data were entered in an Ms Excel spreadsheet and analysed using SPSS statistical Package version 21. Descriptive statistics and chi square tests were performed.

## Results

189 families with children with developmental disabilities were recruited into the study. The mean ± SD age at evaluation of children with developmental disabilities was 7.7 ± 3.7 years, ranging from 1.5 years to 19 years. The most common diagnosis among children with developmental delay was global developmental delay/ intellectual development disorder (43.4%) followed by autism spectrum disorder (15.9%), language disorder (13.8%), cerebral palsy (13.8%). No psychiatric co morbities were reported. All children who were diagnosed with autism spectrum disorder (ASD) were low functioning.

The diagnosis of the pro bands was summarised in table 1. In total, these children had 114 typically developing elder siblings and 50 typically developing younger sibling. The mean ± SD of their elder sibling was 13.5 ± 5 years and that of their younger sibling was 5 ± 4 years.

**Table 1.** - Sociodemographic characteristics of study participants

| Characteristic | Frequency (%) |
| --- | --- |
| Paternal Occupation |  |
| Unemployed | 1 (0.5) |
| Unskilled | 45 (23.8) |
| Semi-skilled | 3 (1.6) |
| Skilled | 24 (12.7) |
| Clerical | 98 (51.9) |
| Semi-profession | 8 (4.2) |
| Profession | 3 (1.6) |
| Data NA | 7 (3.7) |
| Maternal Education |  |
| Primary | 2 (1.1) |
| Middle | 20 (10.6) |
| High | 68 (36) |
| Intermediate | 50 (26.5) |
| Graduate | 48 (25.4) |
| Professional | 1 (0.5) |
| Religion |  |
| Hindu | 65 (34.4) |
| Muslim | 115 (60.8) |
| Christian | 9 (4.8) |

\*Descriptive statistics
NA- Not Available

The mean ± SD of their father’s age was 39 ± 6 years and that of their mother’s age was 32 ± 6 years. Around 52.4% of the mothers were educated above high school level. The most common paternal occupation was clerical job (51.9%) followed by unskilled works (23.8%). The parental socio occupational data was summarised in table 2.

**Table 2.** - Diagnosis of the probands recruited into the study

| Diagnosis | N | Percentage |
| --- | --- | --- |
| GDD/IDD | 82 | 43.4 |
| ASD | 30 | 15.9 |
| ADHD | 4 | 2.1 |
| Language Disorder | 26 | 13.8 |
| Specific Learning Disorder | 6 | 3.2 |
| Cerebral Palsy | 26 | 13.8 |
| Visual Impairment | 2 | 1.1 |
| Hearing Impairment | 2 | 1.1 |
| Diagnosis data NA | 11 | 5.8 |

GDD- Global Developmental Delay; IDD- Intellectual Development Disorder; ASD- Autism Spectrum Disorder; ADHD- Attention Deficit Hyperactivity Disorder; NA- Not Available
\*Descriptive statistics

The proportion of overall complete vaccination among children with developmental disabilities group (70%) and the younger sibling group (64%) were significantly lower than their older sibling group (85%, Pearson chi-square, P<0.01) (see figure 1). The proportion of MMR vaccination among children with developmental disabilities group (60%) and the younger sibling group (59%) were significantly lower than their older sibling group (86%, Pearson chi-square, P<0.001) (see figure 2). There were no significant difference in the proportion of overall vaccination status and MMR vaccination status between index and younger sibling. There was also no association found between vaccination status and sociodemographic variables such as parental education, paternal occupation and religious beliefs.

**Figure 1.**
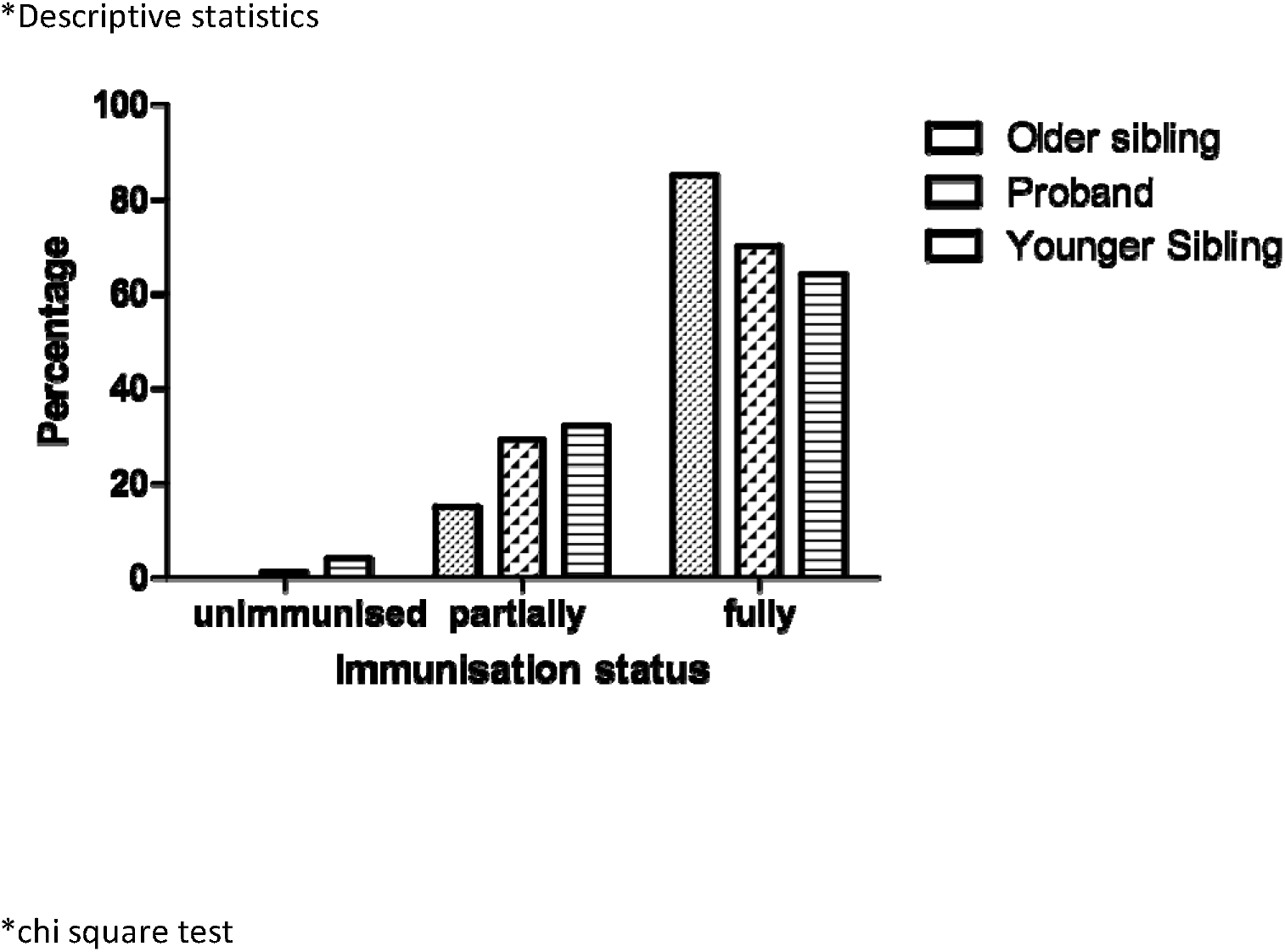
Vaccination status among the study participants

**Figure 2.**
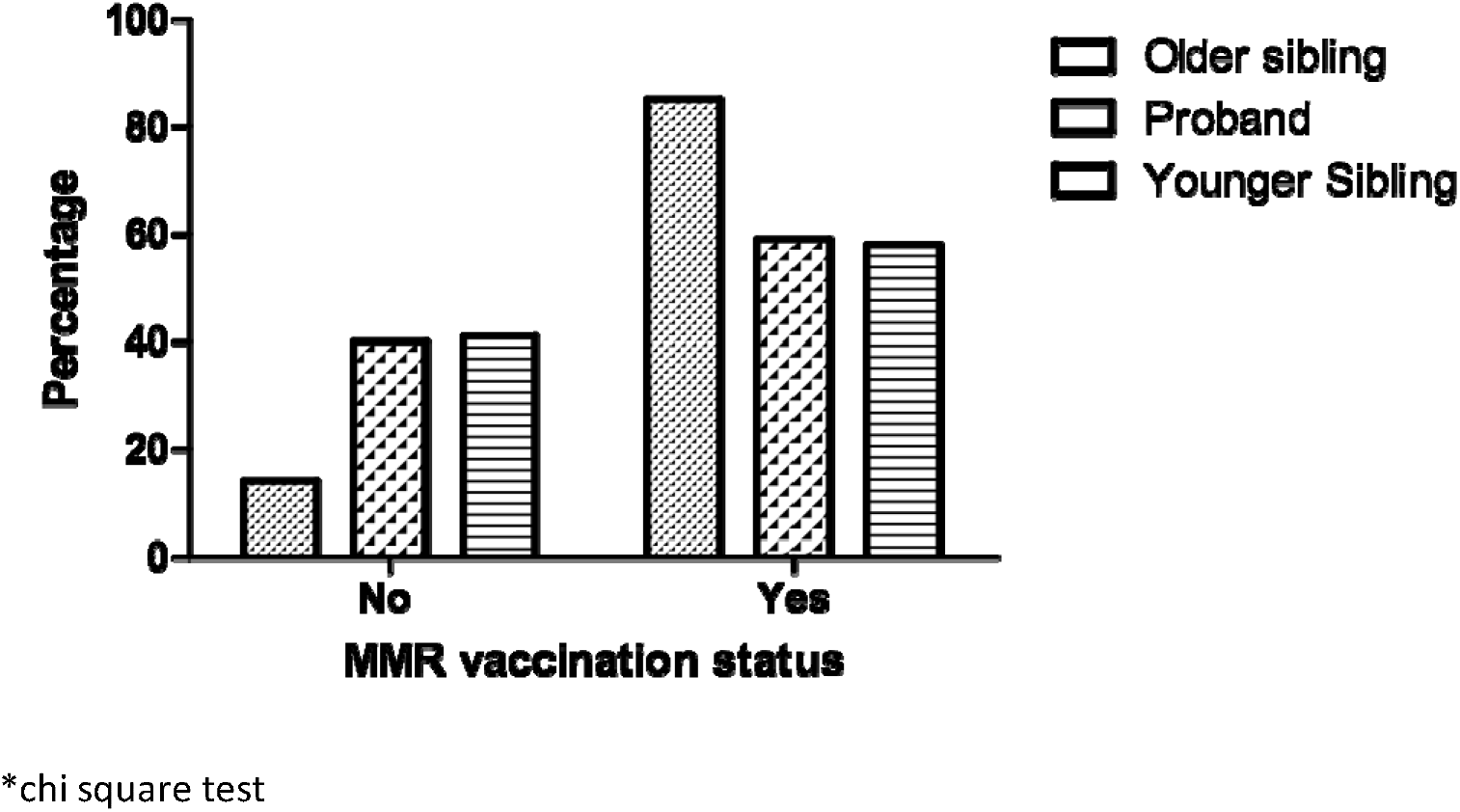
MMR vaccination status among study participants

## Discussion

In our study, parents of children with developmental delay have incompletely vaccinated their children with developmental delay and their younger siblings, resulting in significantly lower immunization rates in probands and later-born children than their older sibling without developmental delay. To the best of our knowledge, this is the first study that have made an attempt to compare vaccination behavior between affected children and their younger sibling to that for their older child without developmental delay. Our findings have important implications for reduced herd immunity and increased vulnerability to vaccine-preventable illness in this high-risk group.

Our study results showed a significantly lower complete vaccination uptake among children with developmental delay when compared with their older siblings. Although parents in the present study were not explicitly asked the reasons for incomplete immunizations, it could be related to both parental and their primary care physicians attitude. There are reports of a supportive role of primary care physicians in delaying immunization in vulnerable children like children with developmental disabilities which could have reinforced the parental concern while attending immunization visit. A meta analysis investigating factors associated with vaccination of young children found that perceived adverse effects of the vaccine and vaccine recommendations by health professionals impact parent’s behaviour.^[3]^

Unlike our study, another similar study on vaccination uptake among sibling of children with ASD reported that the drop in vaccination rate only in younger sibling and not in the index case. This may be due to the fact that diagnosis of ASD may be delayed when compared to diagnosis of other developmental disorders. Thus, this delay in diagnosis of ASD might be the reason the vaccination rate did not decrease in the cases.^[6]^

Speculation that measles, mumps, and rubella (MMR) vaccine may cause autism has been one of the most damaging controversies in vaccine safety which was based on a report in The Lancet in 1998.^[7]^ Even though multiple studies latter found a negative association, some parents still believe this idea leading to the renewed outbreaks of measles in the United States and resurgence of measles in Europe.^[8,9,10]^ Another controversy was regarding the mercury-containing preservative thimerosal in the vaccine which has also been feared to possibly increase the risk of autism. However, a meta-analysis of several epidemiologic studies found no increased risk of autism associated with thimerosal-containing vaccines.^[11]^ Other controversies were related to the association between vaccination and Guillain-Barré syndrome (GBS), its ability to cause chronic autoimmune diseases, and the association of aluminum adjuvants and macrophagic myofasciitis, However, studies failed to prove any of this associations.^[12]^

Our study results also showed a significantly lower MMR vaccination among siblings of children with developmental delay when compared to their older siblings. This may indicate that the parental anxiety regarding the possible association between immunization and autistic spectrum disorders may be having a spillover effect on vaccination behavior of families with children with developmental disabilities in general. This fear has been raising since mid-1990 following a study which argued that the MMR vaccine may lead to the development of autism spectrum disorder.^[7]^ This is despite multiple evidence to the contrary. Studies have also found an equal incidence and age at diagnosis of autism in vaccinated and unvaccinated children.^[8]^ Furthermore, epidemiological data found no influence of MMR on the increased rate of reporting of autism to the General Practice Research database.^[13]^

Our study has the following limitations. The methodology was cross sectional and clinical note based done at developmental centres. No clinical evaluation of the study participants were done by psychiatrists to validate the recorded diagnosis in the clinical note. Moreover, absence of a normal control also limit our interpretation of the study results. A prospective evaluation in a community sample with the involvement of psychiatrists in clarifying the diagnosis and with a control group of normal children could be done in the future for better understanding of vaccination behaviour in this vulnerable group. Moreover, no policy recommendations were possible based on the present study as it was a survey study. Further studies will be required to confirm the findings of this study to take this further for policy recommendations.

## Conclusion

Findings from this study suggest that the reduced vaccination uptake is a trend in families of children with developmental delay attending developmental centres in North Kerala. Previous studies in autism had reported the decline in vaccination uptake only in the younger siblings and not in the proband with autism. In our study, the reduced vaccination uptake was seen both in the proband (children with developmental delay) and their younger sibling. Such a fall in the vaccination rate in this group of children attending developmental centres will make them vulnerable in case of outbreaks. This emphasise the need to have public health strategies targeted to improve the vaccination rate in families of children with developmental disabilities.

## Data Availability

All data related to the manyscript are available

## References

1. World Health Organization. Children: reducing mortality. Fact sheet. <http://www.who.int/mediacentre/factsheets/fs178/en>; 2016.

2. Rainey JJ, Watkins M, Ryman TK, Sandhu P, Bo A, Banerjee K. Reasons related to non-vaccination and under-vaccination of children in low and middle income countries: findings from a systematic review of the published literature, 1999–2009. Vaccine 2011;29:8215–21.

3. Glickman G, Harrison E, Dobkins K. Vaccination Rates among Younger Siblings of Children with Autism. New England Journal of Medicine 2017;377(11):1099–1101. https://doi.org/10.1056/NEJMc1708223

4. Kuwaik G, Roberts W, Brian J, Bryson S, Smith IM, Szatmari P. Zwaigenbaum L. Immunization uptake in siblings of children with autism. Pediatrics 2008;122(3):684–5-6. https://doi.org/10.1542/peds.2008-1624.

5. Smith LE, Amlôt R, Weinman J, Yiend J, Rubin GJ. A systematic review of factors affecting vaccine uptake in young children. Vaccine 2017;35: 6059-6069. pmid:28974409.

6. Abu Kuwaik G, Roberts W, Zwaigenbaum L, Bryson S, Smith IM, Szatmari P, Modi BM, Tanel N, Brian J. Immunization uptake in younger siblings of children with autism spectrum disorder. Autism 2014; 18(2): 148–55. doi: 10.1177/1362361312459111.

7. Wakefield AJ, Murch SH, Anthony A, Linnell J, Casson DM, Malik M, Berelowitz M, Dhillon AP, Thomson MA, Harvey P, Valentine A, Davies SE, Walker-Smith JA. Ileal-lymphoid-nodu-lar hyperplasia, nonspecific colitis, and pervasive developmental disorder in children. Lancet 1998;351:637–41.

8. Taylor B, Miller E, Farrington CP, et al. Autism and measles, mumps, and rubella vaccine: no epidemiological evidence for a causal association. Lancet 1999;353:2026–9.

9. Madsen KM, Hviid A, Vestergaard M, et al. A population-based study of measles, mumps, and rubella vaccination and autism. N Engl J Med 2002;347:1477–82.

10. Smeeth L, Cook C, Fombonne E, et al. MMR vaccination and pervasive developmental disorders: a case-control study. Lancet 2004; 364: 963–9.

11. Taylor LE, Swerdfeger AL, Eslick GD. Vaccines are not associated with autism: an evidence-based meta-analysis of case-control and cohort studies. Vaccine 2014;32:3623–9.

12. DeStefano F, Bodenstab HM, Offit PA. Principal controversies in vaccine safety in the United States. Clinical Infectious Diseases 2019;69(4):726–31.

13. Farrington CP, Miller E, Taylor B. MMR and autism: further evidence against a causal association. Vaccine 2001; 19(27):3632–5.

